# Application of a novel analytical pipeline for high-dimensional multivariate mediation analysis of environmental data

**DOI:** 10.1101/2020.05.30.20117655

**Authors:** Max T. Aung, Yanyi Song, Kelly K. Ferguson, David E. Cantonwine, Lixia Zeng, Thomas F. McElrath, Subramaniam Pennathur, John D. Meeker, Bhramar Mukherjee

**Author notes:** Corresponding Author: Bhramar Mukherjee, University of Michigan School of Public Health, Department of Biostatistics, M4208 SPH II, 1415 Washington Heights Ann Arbor, Michigan 48109-2029.

## Abstract

Diverse toxicological mechanisms may mediate the impact of environmental toxicants (phthalates, phenols, polycyclic aromatic hydrocarbons, and metals) on pregnancy outcomes. In this study we introduce an analytical pipeline for high-dimensional mediation analysis to identify mediation pathways (*q* = 63 mediators) in the relationship between environmental toxicants (*p* = 38 analytes) and gestational age at delivery. Our analytical pipeline included: (1) conducting pairwise mediation for unique exposure-mediator combinations, (2) subjecting mediators to Bayesian shrinkage mediation analysis and population value decomposition, and (3) exposure dimension reduction by estimating environmental risk scores. Dimension reduction demonstrated that a one unit increase in phthalate risk score was associated with a total effect of 1.09 lower gestational age (in weeks) at delivery (95% confidence interval: 1.78 – 0.36) and eicosanoids from the cytochrome p450 pathway mediated 24.5% of this effect (95% confidence interval: 4%-66%). Eicosanoid products derived from the cytochrome p450 pathway may be important mediators of phthalate toxicity.

## 1. Introduction

Globally, humans are susceptible to widespread exposure to environmental contamination through several anthropogenic operations, including manufacturing of industrial chemicals, natural resource extraction, and fossil fuel combustion. Pregnant women and developing fetuses are uniquely vulnerable to environmental exposures. The prenatal exposome encompasses the totality of environmental exposures the occur preconception and during pregnancy. Among the vast landscape of pollutants that exist in modern society, we focus the attention of this present study on four classes of environmental contaminants: phthalates, phenols, polycyclic aromatic hydrocarbons, and trace metals. Phthalates are high-production volume chemicals used as plasticizers in numerous consumer products such as vinyl flooring, children’s toys, and food packaging^1^. Phenol derived compounds are also widely used in plastics, as well as personal care and pharmaceutical products^2,3^. Polycyclic aromatic hydrocarbons commonly enter the environment through industrial and natural processes, such as combustion of coal, fossil fuel, and organic debris^4^. Additional anthropogenic activities such as natural resource extraction and electronic waste recycling can yield environmental contamination of heavy metals (e.g. lead, cadmium, mercury)^5^. Biomonitoring analytes derived from these toxicant classes can capture the magnitude of exposure in humans to inform health studies.

Exposures to environmental toxicant classes are suspected risk factors for adverse pregnancy outcomes such as preterm delivery or shortened gestational duration^6^. Based on extensive *in vitro* and animal models, there is evidence indicating inducible receptor activity attributable to toxicant classes (e.g. phthalates and phenols with the estrogen receptor, polycyclic aromatic hydrocarbons and the aryl hydrocarbon receptor, toxic heavy metals and NF-κB)^7,8^. Perturbations in receptor signaling and transcription factor activity can result in altered production of signaling molecules responsible for inflammation and metabolism. Therefore, biomarker signals of toxicant exposures can help disentangle the mediating biological pathways linking toxicant mixtures to pregnancy outcomes, which is critical for early detection of disease and prevention. Mediation pathways can also inform causal inference in human observational studies and thus the need for policy driven exposure reduction and prevention measures.

In previous analyses in the LIFECODES cohort, we have found that biomarkers of inflammation (cytokines and c-reactive protein [CRP]) and oxidative stress (8-isoprostane) have been associated with increased risk of preterm birth^9,10^. Additionally, we learned that cytochrome p450 and lipoxygenase derived eicosanoids had the greatest relative predictive capacity for classifying preterm birth compared to a large panel of endogenous biomarkers^11^. Eicosanoids are potent signaling molecules derived from parent fatty acid compounds such as arachidonic acid and linoleic acid^12^. For example, epoxyeicosatrienoic acids (EETs) produced through the cytochrome p450 pathways are involved in vascular remodeling and regulation of inflammation^13^. Therefore these eicosanoids may be up-regulated in disease states involving cardiovascular irregularities, an important precursor for adverse pregnancy outcomes^14^. Among the environmental exposures, we also showed that phthalates have been associated with increased risk for preterm delivery and reduced gestational age at delivery in the larger case-control study within the LIFECODES cohort^15,16^. Based on these previous studies, there is circumstantial evidence of potential mediation between prenatal phthalate exposure and pregnancy outcomes through eicosanoids.

Precise risk profiles can be estimated through various inferential tools, including the implementation of causal mediation analysis. Early methodological development of mediation analysis enabled investigators to decompose the relationship between a predictor and an outcome, and test whether the total effect is partially mediated by an intermediate variable^17,18^. With the expansion of technology to measure increasingly high-dimensional endogenous biomarkers, the mediation framework needs to evolve in order handle the analysis of multiple mediators simultaneously. Theoretical advancements in the mediation analysis framework have afforded new applications of mediation analysis to account for multiple mediators. For example, Chen and colleagues have developed methodological applications for leveraging population value decomposition to reduce the mediator matrix and analyze multiple mediators simultaneously^19^. More recently, Song and colleagues developed a Bayesian approach for continuous shrinkage estimation of multiple predictors to identify the most important mediation pathways^20^. In another study, Zhang and colleagues sought to approach this high-dimensional mediation scenario through penalized regularization using the minimax concave penalty and joint significance testing^21^. Another regularization approach that has been applied is a novel lasso penalty on mediating pathways^22^. Altogether, these statistical methods have pushed the frontier of mediation analysis and created opportunities to make critical scientific discoveries using biomarkers from complex biological pathways.

Currently, there is a critical knowledge gap for addressing a multivariate mediation setting where there are not only high-dimensional mediators, but also multiple toxicants with inherent collinearity and grouping structures. Our study aims to advance methodological applications to investigate a mixtures mediation setting with multiple toxicant classes and groups of endogenous biomarker mediators. We propose a novel analytical pipeline that integrates our previous methodological contributions in environmental risk score construction^16,23,24^ with the aforementioned multivariate mediation analysis methods^19,21^. In this pipeline, we provide a guided step-wise approach that begins with one-at-a-time pairwise mediation, followed by mediator shrinkage and dimension reduction, and arrives at mediation analysis with both exposure and mediator dimension reduction.

## 2. Results

The weighted characteristics of our study sample are reported in **Table 1**. Our study sample was predominantly White (65.7%) and college educated (79.2%). The weighted median age (in years) of participants was 32.8 (interquartile range [IQR]: 4.8). The weighted median gestational age at delivery (in weeks) across the study sample was 38.7 (IQR: 2.0). Overall, a majority (51.6%) of participants had a BMI < 25kg/m^2^ at their initial visit. There were very few participants who smoked (5.4%) or drank alcohol (5.0%) during pregnancy.

**Table 1.**
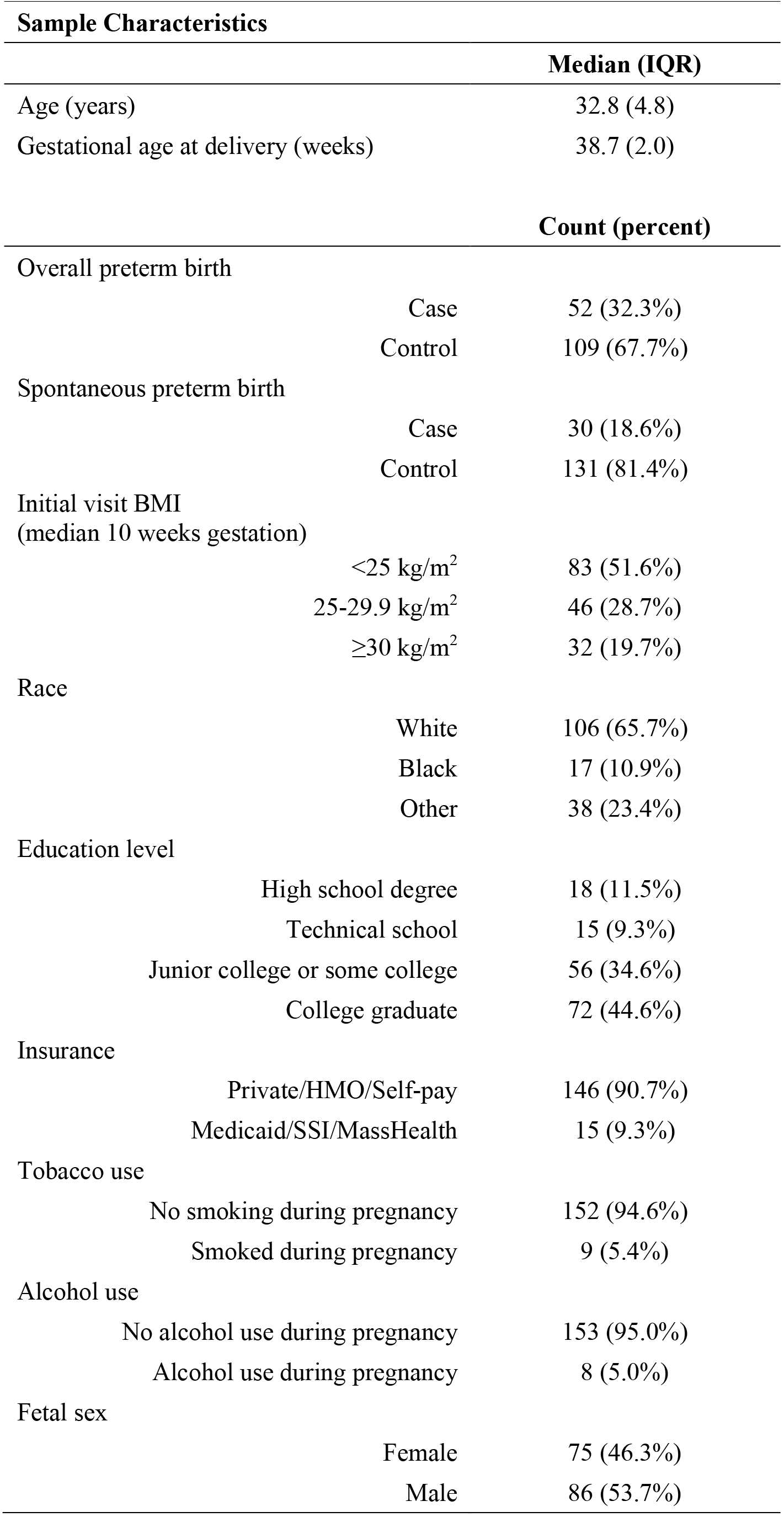
LIFECODES Prospective pregnancy cohort profile, weighted for inverse probability weights of case-control sampling (N = 161)

| Sample Characteristics |  |  |
| --- | --- | --- |
|  |  | Median (IQR) |
| Age (years) |  | 32.8 (4.8) |
| Gestational age at delivery (weeks) |  | 38.7 (2.0) |
|  |  | Count (percent) |
| Overall preterm birth |  |  |
| Case |  | 52 (32.3%) |
| Control |  | 109 (67.7%) |
| Spontaneous preterm birth |  |  |
| Case |  | 30 (18.6%) |
| Control |  | 131 (81.4%) |
| Initial visit BMI<br>(median 10 weeks gestation) |  |  |
| <25 kg/m<sup>2</sup> |  | 83 (51.6%) |
| 25-29.9 kg/m<sup>2</sup> |  | 46 (28.7%) |
| ≥30 kg/m<sup>2</sup> |  | 32 (19.7%) |
| Race |  |  |
| White |  | 106 (65.7%) |
| Black |  | 17 (10.9%) |
| Other |  | 38 (23.4%) |
| Education level |  |  |
| High school degree |  | 18 (11.5%) |
| Technical school |  | 15 (9.3%) |
| Junior college or some college |  | 56 (34.6%) |
| College graduate |  | 72 (44.6%) |
| Insurance |  |  |
| Private/HMO/Self-pay |  | 146 (90.7%) |
| Medicaid/SSI/MassHealth |  | 15 (9.3%) |
| Tobacco use |  |  |
| No smoking during pregnancy |  | 152 (94.6%) |
| Smoked during pregnancy |  | 9 (5.4%) |
| Alcohol use |  |  |
| No alcohol use during pregnancy |  | 153 (95.0%) |
| Alcohol use during pregnancy |  | 8 (5.0%) |
| Fetal sex |  |  |
| Female |  | 75 (46.3%) |
| Male |  | 86 (53.7%) |

### 2.1 Associations between exposures and outcomes

We present forest plots of single pollutant associations with overall preterm birth (Supplemental **Figure 1**), spontaneous preterm birth (Supplemental **Figure 2**), and gestational age at delivery (**Figure 2**). Given the limited power in sample size for this study, we observed positive associations between phthalate metabolites (Mono-2-ethyl-hexyl phthalate [MEHP], mono-2-ethyl-5-hydroxyhexyl phthalate [MEHHP], mono-2-ethyl-5-oxohexyl phthalate [MEOHP], and mono-2-ethyl-5-carboxypentyl phthalate [MECPP]) and odds of overall preterm birth and spontaneous preterm birth, however these estimates were not significantly different from the null. Associations with phthalates have been previously explored in the larger LIFECODES cohort, where several of theses associations were significant^15,16^. Three toxicants (2,4-dichlorophenol [2,4-DCP], 1-hydroxyphenanthrene [1-PHE], and thallium [Tl]) were associated with decreased odds of overall preterm birth (**Figure 2**). When focusing on spontaneous preterm birth, five toxicants (2,4-DCP, 2,5-dichlorophenol [2,5-DCP], triclosan [TCS], 9-hydroxyphenanthrene [9-PHE], and 1-PHE) were associated with decreased odds (Supplemental **Figure 2**). Similar directions of associations were observed in a previous analysis of these phenols in the larger LIFECODES cohort as well^25^. Expectedly, modeling gestational age at delivery revealed that the same phthalate metabolites (MEHP, MEHHP, MEOHP, and MECPP) that were suggestively associated with preterm birth were also suggestively associated with decreased gestational age at delivery (**Figure 2**). Copper [Cu] was also associated with shortened gestational age at delivery, while 2,5-DCP and molybdenum [Mo] were associated with increased gestational age at delivery (**Figure 2**). We also observed consistent findings with Cu in a previous analysis in the larger LIFECODES cohort, where Cu was associated with increased odds of overall preterm birth^26^.

**Figure 1.**
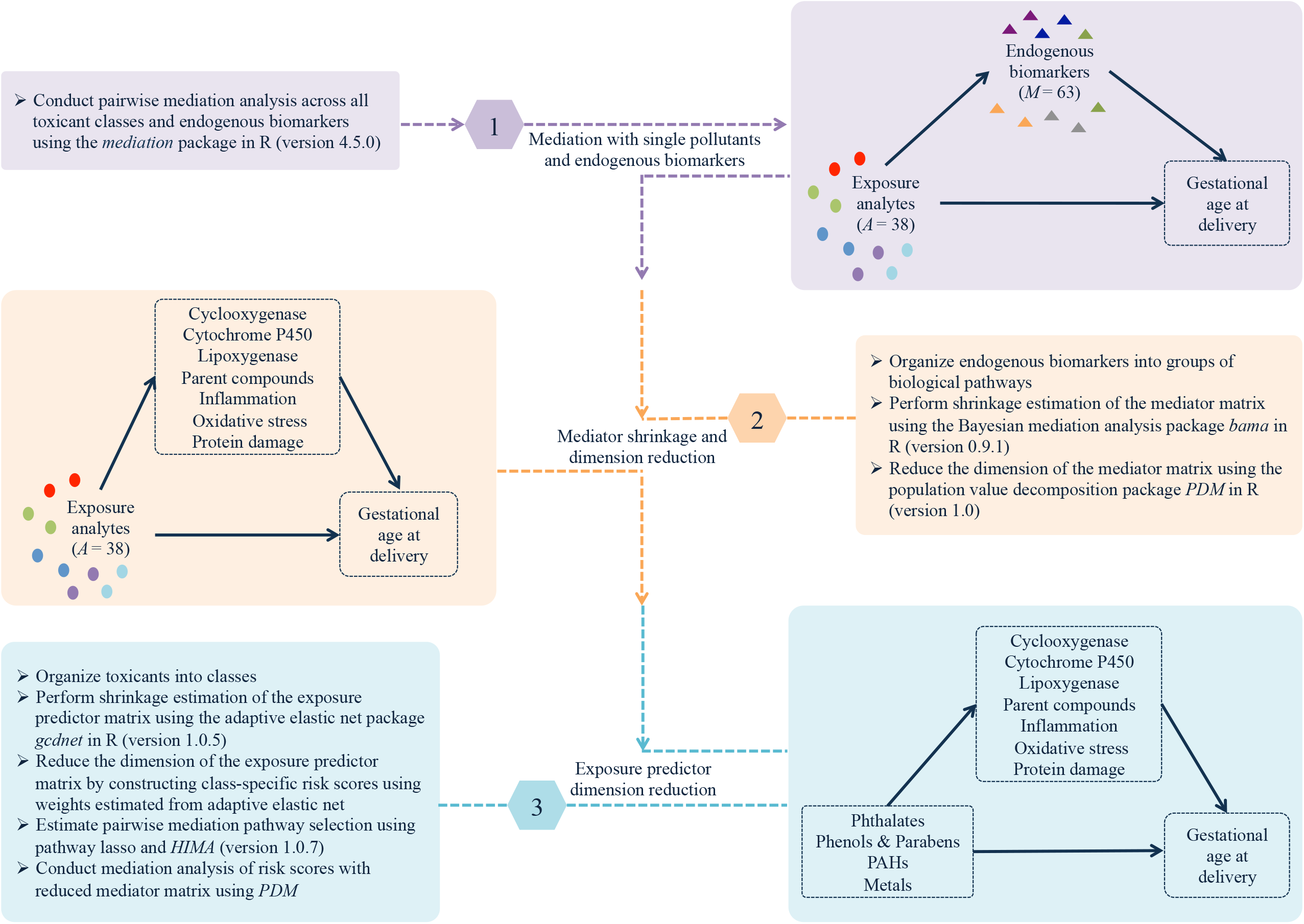
Analytical pipeline for conducting high-dimensional mediation analysis of toxicant mixtures, endogenous biomarkers, and birth outcomes.

**Figure 2.**
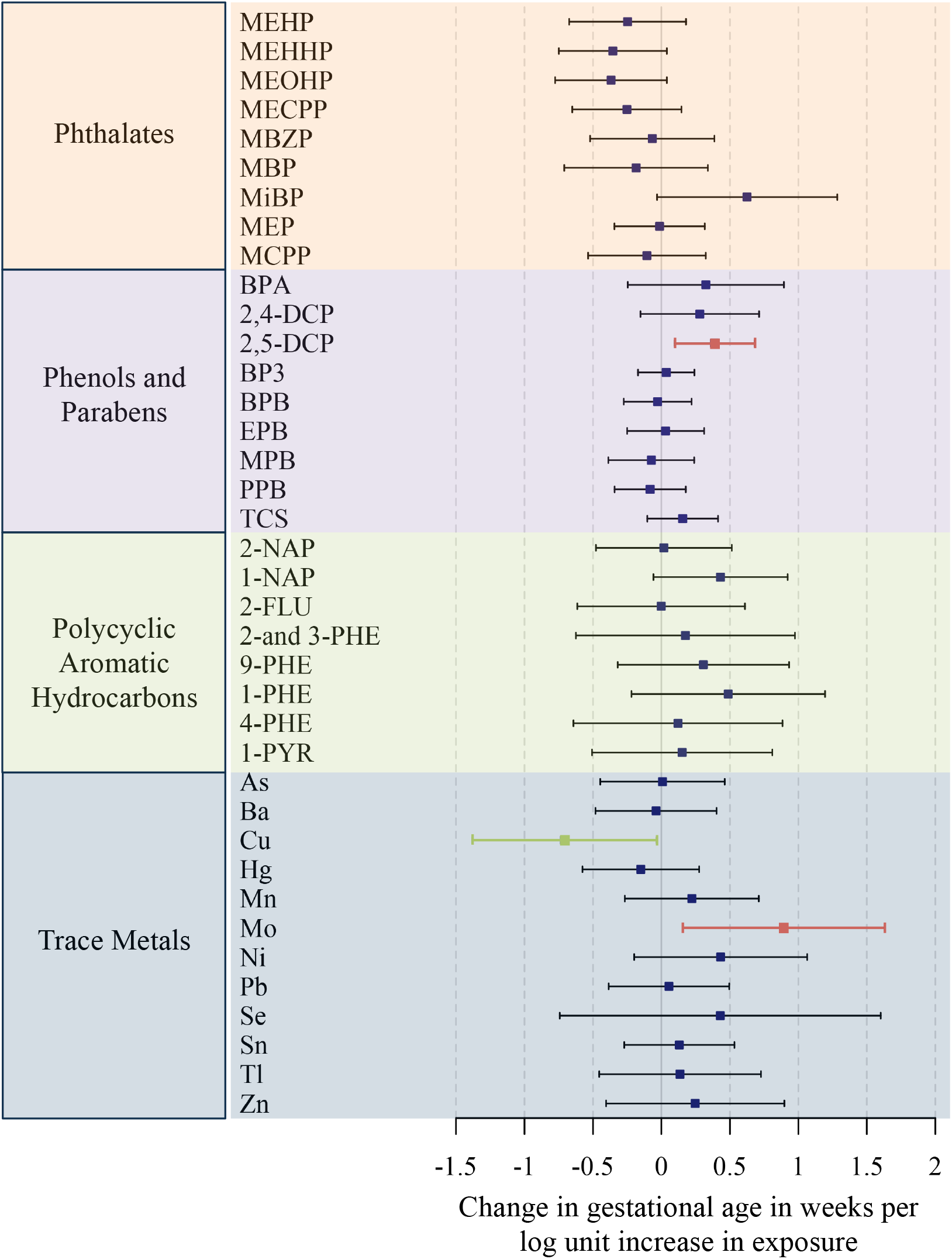
Associations between individual toxicants and gestational age at delivery (in weeks) estimated by weighted multiple linear regression (N = 161). Models adjusted for specific gravity, maternal age, race, and BMI at initial study visit.

### 2.2 Pairwise mediation

We present –log_10_(p-values) for the estimated natural indirect effects for each pair of toxicants and mediators for models of overall preterm birth (Supplemental **Figure 3**), spontaneous preterm birth (Supplemental **Figure 4**), and gestational age at delivery (**Figure 3**). For overall preterm birth, we observed 16 natural indirect effects (p< 0.05), although their corresponding direct and total effects were not significant (Supplemental **Figure 3** and **Supplemental Table 1.A**). The mediators that appeared the most among these mediation effects were resolvin D1 [RVD1] and 11,12-dihydroxy-eicosatrienoic acid [11,12-DHET] (Supplemental **Figure 3**). There were 56 natural indirect effects (p< 0.05) that were observed for spontaneous preterm birth (Supplemental **Figure 4**). Among those mediation pathways for spontaneous preterm birth, one pair (TCS and 11,12-DHET) also exhibited significant total and direct effects, where 11,12-DHET mediated 19.1% of the protective total effect of TCS on spontaneous preterm birth (**Supplemental Table 1.B**). Last, there were five natural indirect effects (p< 0.05) observed for gestational age at delivery, however the total and direct effects were not significant (**Figure 3** and **Supplemental Table 1.C**).

**Figure 3.**
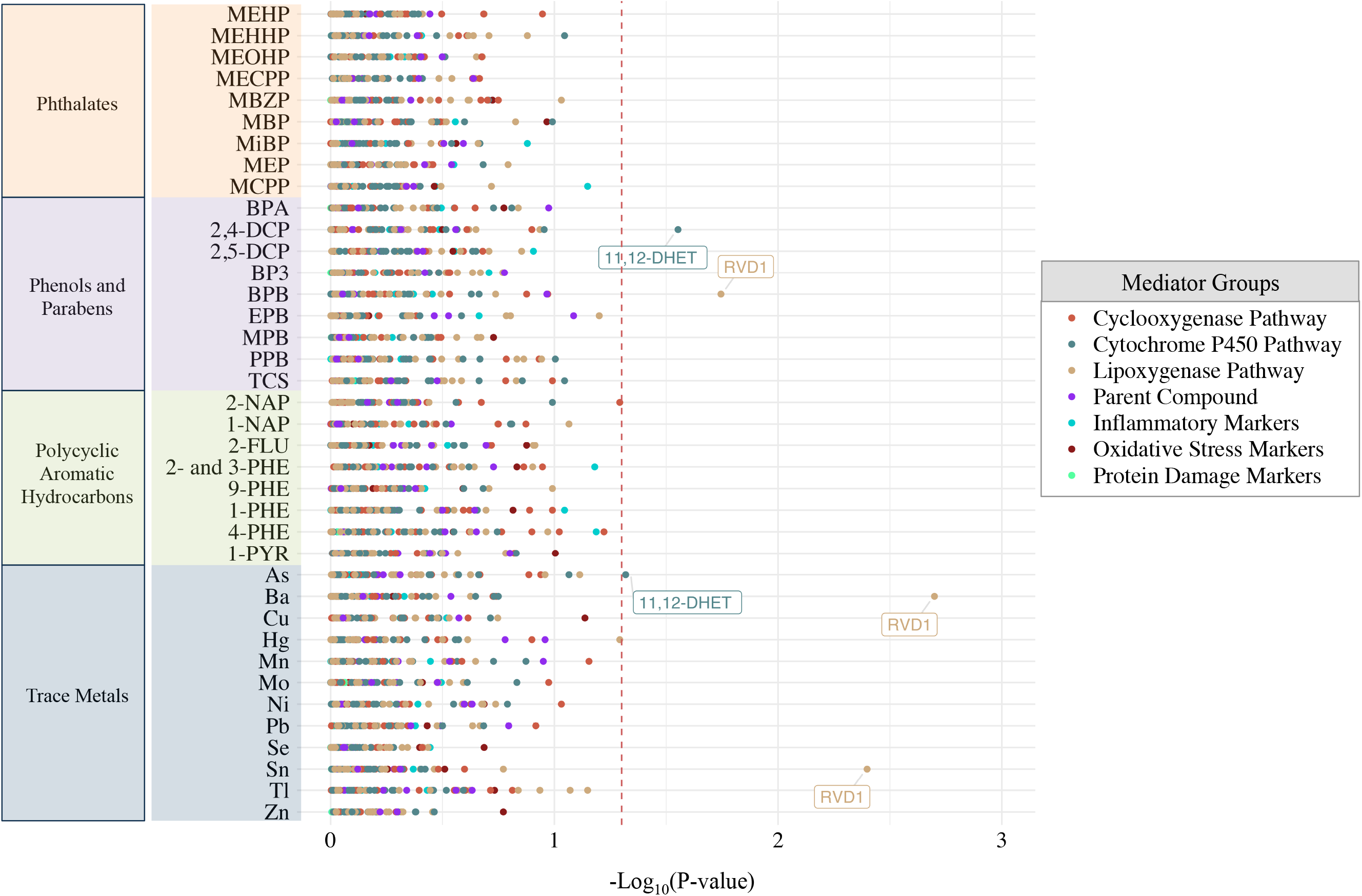
Estimated −log_10_(p-values) of natural indirect effects from pairwise weighted mediation models for gestational age at delivery (N = 161). Dashed vertical red line signifies p-value threshold of 0.05, and labeled point estimates are below that threshold. Models adjusted for specific gravity, maternal age, race, and BMI at initial study visit.

**Figure 4.**
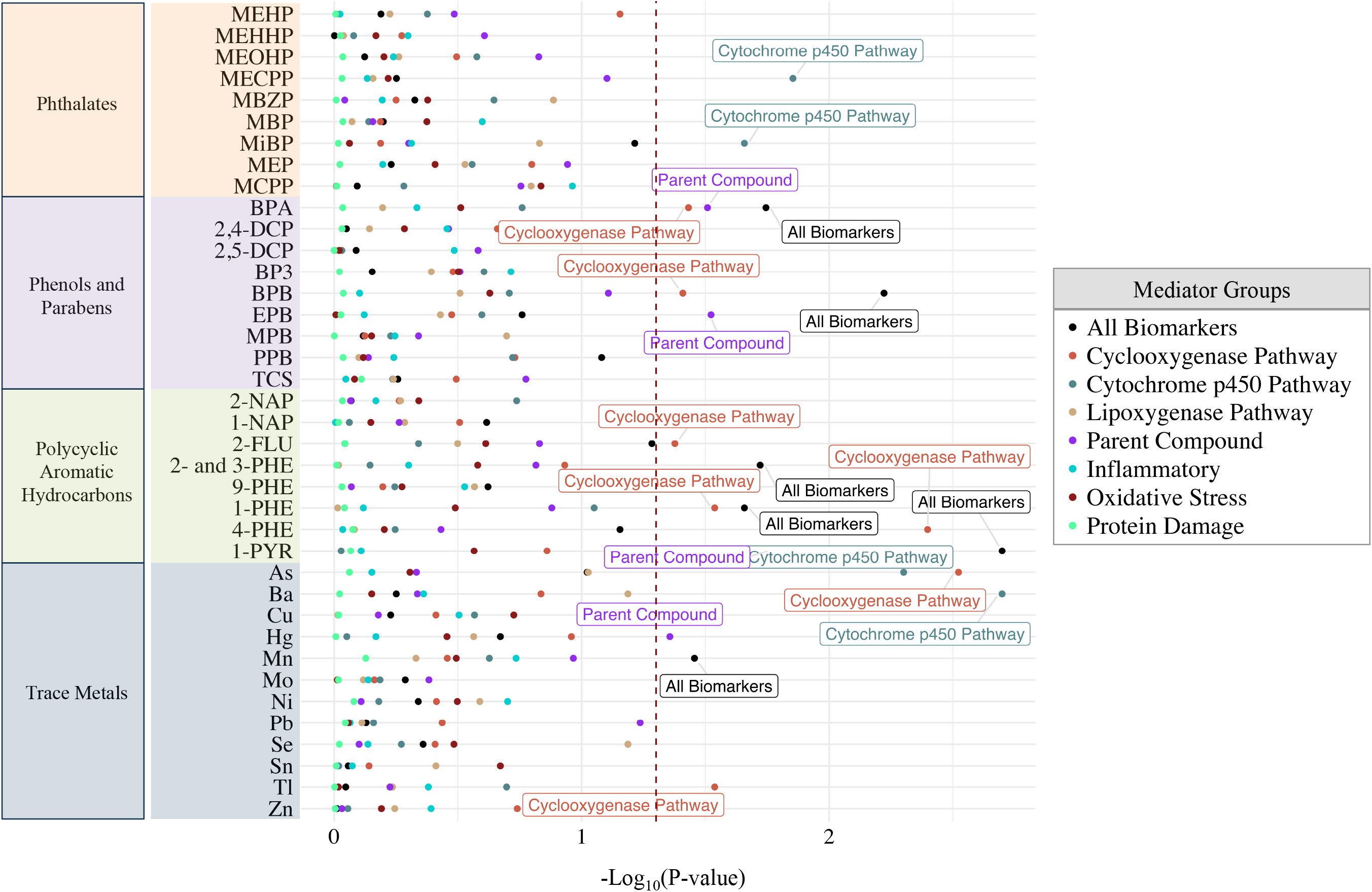
Estimated –log_10_(p-values) of natural indirect effects from pairwise mediation models for gestational age at delivery (N = 161) using mediator group effects derived from population value decomposition. Dashed vertical red line signifies p-value threshold of 0.05, and labeled point estimates are below that threshold. Models adjusted for specific gravity, maternal age, race, and BMI at initial study visit.

### 2.3 Multiple mediators: shrinkage and dimension reduction

Bayesian continuous mediation shrinkage and population value decomposition for mediator dimension reduction were both restricted to the continuous outcome variable gestational age at delivery. When we conducted mediator shrinkage, mediation pathway signals were sparse and posterior inclusion probabilities did not exceed 0.1 (**Supplemental Table 2**). Mediator dimension reduction allowed us to evaluate whole groups of mediators as potential mediation pathways for individual toxicants. In this approach, we observed natural indirect effects (p< 0.05) for MECPP and mono-isobutyl phthalate (MiBP) with the cytochrome p450 pathway (**Figure 4**). MECPP in particular had a positive indirect effect (p< 0.05) and suggestively positive (p< 0.1) total effect, however the direction of the natural indirect effect was negative – indicating inconsistent mediation (**Supplemental table 3**). Multiple phenols demonstrated natural indirect effects with parent lipid compounds (bisphenol-A [BPA] and ethyl paraben [EPB]) and the cyclooxygenase pathway (BPA and butyl paraben [BPB]), however these mediation pathways did not demonstrate significant total and direct effects (**Figure 4** and **Supplemental Table 3**). Similarly, three polycyclic aromatic hydrocarbons (1-PHE, 2-hydroxyfluorene [2-FLU], and 4-hydroxyphenanthrene [4-PHE]) demonstrated natural indirect effects with cyclooxygenase pathway, however their total and direct effects were not significant (**Figure 4** and **Supplemental Table 3**). Among the trace metals, arsenic [As] and barium [Ba] exhibited natural indirect effects for the cytochrome p450 pathway without total or direct effects (**Figure 4** and **Supplemental Table 3**). We also observed natural indirect effects for the cyclooxygenase pathway attributable to As and thallium [Tl], albeit the total and direct effects were not significant (**Figure 4** and **Supplemental Table 3**).

### 2.4 Both exposure and mediator dimension reduction

Continuing to focus strictly on gestational age at delivery, we reduced the dimensionality of the exposure matrix by creating environmental risk scores for each toxicant class: phthalates, phenols, metals, and polycyclic aromatic hydrocarbons. **Figure 5** illustrates forest plots of total effects, direct effects, and natural indirect effects for each environmental risk score across each mediator group (All biomarkers, cyclooxygenase pathway, cytochrome p450 pathway, lipoxygenase pathway, parent lipid compounds, inflammatory biomarkers, oxidative stress biomarkers, and protein damage biomarkers). The phthalate risk score exhibited natural indirect effects among all biomarkers, which was chiefly driven by the cytochrome p450 pathway (**Figure 5**). One unit increase in the phthalate risk score was associated with a total effect of 1.09 lower gestational age (in weeks) at delivery (95% confidence interval: 1.78 – 0.36) (**Figure 5** and **Supplemental Table 4**). From this total effect, 24.5% of the effect was mediated through the cytochrome p450 pathway (**Figure 5** and **Supplemental Table 4**).

**Figure 5.**
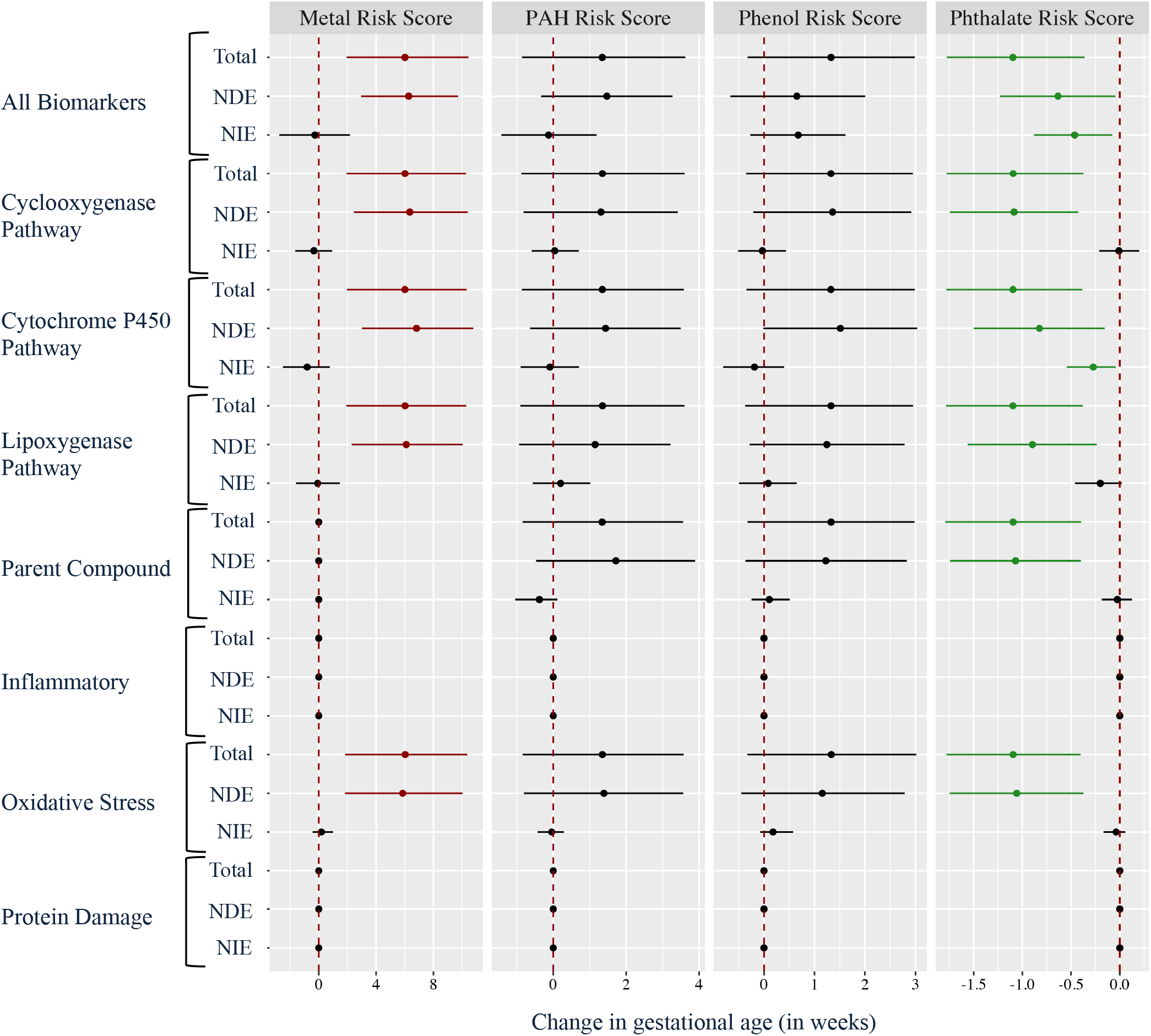
Forest plot of mediation analysis for environmental risk scores, mediator group effects, and gestational age at delivery. Models adjusted for specific gravity, maternal age, race, and BMI at initial study visit.

Abbreviations: Natural direct effect (NDE); Natural indirect effect (NIE)

**Table 2** reports mediators that remained for each risk score after applying pathway lasso shrinkage. We observed that select mediators from the cytochrome p450 (17-hydroxy-eicosatetraenoic acid [17-HETE], 18-hydroxy-eicosatetraenoic acid [18-HETE]), lipoxygenase pathway (Leukotriene B4 [LTB4]), parent lipid compounds (Docosahexaenoic acid [DHA]), inflammation (CRP, interleukin-1β [IL-1β], interleukin-6 [IL-6], interleukin-10 [IL-10], tumor necrosis factor-α [TNF-α]) remained in the relationship between the phthalate risk score and gestational at delivery (**Table 2**). For the metal risk score, pathway lasso selected mediators in the cyclooxygenase pathway (Prostaglandin B2 [PGB2], prostaglandin A2 [PGA2], prostaglandin E1 [PGE1], and bicycle-prostaglandin E2 [BCPGE2]), cytochrome p450 pathway (17-HETE, 18-HETE, and 11,12-DHET), inflammation (IL-6), and protein damage group (CY) (**Table 2**). For the phenol risk score, pathway lasso selected mediators from the cytochrome p450 pathway (17-HETE, 9,10-dihydroxy-octadecenoic acid [9,10-DiHOME], and 8,9-epoxy-eicsatrienoic acid [8(9)-EET]), lipoxygenase pathway (13-oxooctadeca-dienoic acid [13-oxoODE]), inflammation [CRP], and oxidative stress group (8-hydroxydeoxyguanosine [8-OHdG] and 8-isoprostane [8-IP]) (**Table 2**). The polycyclic aromatic hydrocarbon risk score was linked with two mediators, from the cyclooxygenase pathway (PGE1), and oxidative stress group (8-IP) (**Table 2**).

**Table 2.** Pairs of environmental risk scores and mediators selected by pathway lasso.

|  | Metal Risk Score | PAH Risk Score | Phenol Risk Score | Phthalate Risk Score |
| --- | --- | --- | --- | --- |
| Cyclooxygenase Pathway | PGB2, PGA2, PGE1, BCPGE2 | PGE1 | - | - |
| Cytochrome P450 Pathway | 17-HETE, 18-HETE, 11,12-DHET | - | 17-HETE, 9,10-DiHOME, 8(9)-EET | 17-HETE, 18-HETE |
| Lipoxygenase Pathway | - | - | 13-oxoODE | LTB4 |
| Parent Compound | - | - | - | DHA |
| Inflammatory | IL-6 | - | CRP | CRP, IL-1β, IL-6, IL-10, TNF-α |
| Oxidative Stress | - | 8-IP | 8-OHdG, 8-IP | - |
| Protein Damage | CY | - | - | - |

In **Table 3**, we document the findings from the *hima* function that applies a minimax concave penalty and joint significance test for mediator pathways. We observed that in this method, the phthalate risk score was significantly linked (FDR q-value < 0.2) to mediators in the lipoxygenase pathway (RVD1) and inflammation group (IL-10) (**Table 3**). Although not meeting the FDR threshold, select cytochrome p450 products (20-hydroxy-eicosatetraenoic acid [20-HETE], 20-carboxy arachidonic acid [CAA], and 11,12-DHET) also showed subtle mediation signals for the phthalate risk score (**Table 3**). The phenol risk score was linked (FDR q-value < 0.1) to mediators in the cytochrome p450 pathway (20-HETE, and 11,12-DHET) and oxidative stress group (8-OHdG) (**Table 3**). Last, the polycyclic aromatic hydrocarbon risk score was also linked (FDR q-value < 0.2) to the oxidative stress mediator 8-OHdG (**Table 3**).

**Table 3.** Pairs of environmental risk scores and mediators with detected mediation effects estimated using the minimax concave penalty and joint significance test in *hima*.

|  | Metal Risk Score | PAH Risk Score | Phenol Risk Score | Phthalate Risk Score |
| --- | --- | --- | --- | --- |
| Using all biomarkers | IL-10, 8-OHdG, RVD1 | IL-10, 8-OHdG, RVD1 | RVD1 | IL-10**, 8-OHdG, RVD1 |
| Cyclooxygenase Pathway | 13,14DHK-PGD2, PGD3 | PGD3 | PGD3 | - |
| Cytochrome P450 Pathway | 20-HETE, 11,12-DHET | 20-HETE, CAA, 11,12-DHET | 20-HETE**, 11,12-DHET** | 20-HETE, CAA, 11,12-DHET |
| Lipoxygenase Pathway | RVD1 | RVD1 | RVD1 | RVD1* |
| Parent Compound | AA, LA | - | - | - |
| Inflammatory | IL-10 | IL-10 | IL-10 | IL-10** |
| Oxidative Stress | 8-OHdG | 8-OHdG* | 8-OHdG** | 8-OHdG |
| Protein Damage | CY | - | - | - |

\* FDR <0.2
\*\* FDR <0.1

## 3. Discussion

Conducting high-dimensional mediation analysis of toxicant mixtures and endogenous biomarkers advances scientific discovery of intermediate pathways by which environmental contamination may impact pregnancy outcomes. Furthermore, mediation analysis enhances causal inference in observational studies. In this study, we designed a novel pipeline to guide researchers through an important mediation analysis setting that accounts for multiple predictors and mediators. This is particularly applicable for the analysis of high-dimensional data of endogenous omics-scale biomarkers. Our pipeline begins with single pairwise mediation, underlining the sparsity of signals when evaluating any single biomarker. Ultimately, our pipeline arrives towards a dimensionally reduced setting with exposure class risk scores and mediator group effects organized by enzymatic pathways.

Pairwise mediation yielded subtle evidence of natural indirect effects, which was expected given the limited sample size and insufficient power to detect fully significant natural direct and total effects. Nonetheless, the findings from pairwise mediation provide initial evidence of specific toxicant-mediator pairs that may appear in larger study samples. When we reduced the dimensionality of the exposure and mediator matrices, we combined the effects of individual toxicants into their designated classes, therefore estimating their cumulative effect. This approach also combined the mediator contributions into enzymatic pathways or groups. Importantly, the phthalate risk score was associated with decreased gestational age at delivery and this effect was mediated by cytochrome p450 derived eicosanoids. We also observed suggestive evidence of mediation with the lipoxygenase pathway in the association between the phthalate risk score and gestational age at delivery. Overall this analytical pipeline demonstrates the differences observed with single biomarker analysis and whole pathway analysis with toxicant mixtures.

Throughout our analytical pipeline, we explored a sophisticated ensemble of contemporary approaches for high-dimensional mediation analysis: Bayesian continuous shrinkage mediation^20^, population value decomposition^19^, pathway lasso^22^, and application of minimax concave penalty and joint significance testing^21^. The theoretical basis for each of these methods applies unique penalties and estimation algorithms; therefore we expect a substantive degree of non-uniformity in results across methods. Nonetheless, findings that appear repetitively across methods materialize relationships that indicate importance, afford greater confidence of a true mediation pathway, and warrants deeper investigation through replication in independent samples. Chiefly, we learned that the phthalate risk score appears to be linked with the whole cytochrome p450 pathway effect using population value decomposition. When comparing this finding with the other high-dimensional mediation methods, individual cytochrome p450 metabolites were identified in both the pathway lasso penalty and the minimax concave penalty with joint significance testing. Furthermore, there are suggestive signals of lipoxygenase pathway and metabolites being linked to the phthalate risk score across each of these high-dimensional mediation methods. Another important finding was the lack of a mediation effect observed between the phthalate risk score and cyclooxygenase pathway or its metabolites when applying each of the high-dimensional mediation methods. Our study provides consistent evidence that phthalates may impact gestational age at delivery mediated through the cytochrome p450 pathway – and to a lesser degree the lipoxygenase pathway.

The cytochrome p450 enzymes are a conserved superfamily of enzymes that function to metabolize environmental toxicants, drugs, and endogenous compounds^27^. Transformation of endogenous fatty acid compounds such as arachidonic acid results in the production of bioactive signaling molecules, which can influence cellular biology through altering membrane permeability and transcription factor activity^28^. For example, cytochrome p450 derived HETEs can impact renal vascular changes and impact other enzymes, such as inhibiting adenosine triphosphatase [ATPase] activity or stimulating peroxisome proliferator-activated receptor gamma [PPARγ]^29,30^. Additionally, EETs can mediate anti-inflammatory effects by inhibiting NF-κB activity and increasing MAP kinase activity and cyclic AMP concentrations^13^. Cytochrome p450 activity is partially stimulated by elevated pro-inflammatory cytokines such as TNF-α^28^. As such, we expect maternal biomarker profiles to reflect observable differences in cytochrome p450 products in disease states. Furthermore, circulation of cytochrome p450 products is critically important for regulating cardiovascular functions, such as vasodilatory changes and vascular inflammation partly through expression of endothelial adhesion molecules for chemotaxis and leukocytic migration^28^. The underlying mechanisms associated with reduced gestational age duration and preterm delivery include maternal inflammation and systemic oxidative stress^31,32^. Therefore, the cardiovascular and inflammatory responsiveness of cytochrome p450 activity underline the critical importance of these enzymes for pregnancy outcomes.

The mediation signal between phthalates and cytochrome p450 pathway underline an important toxicological mechanism. Several phthalate parent compounds (dimethyl phthalate, diethyl phthalate, dipropyl phthalate, dibutyl phthalate, benzyl butyl phthalate, dicyclohexyl phthalate) have been previously shown to inhibit cytochrome p450 enzymatic activity *in vitro*^33^. However, another *in vitro* study demonstrated that cytochrome p450 activity is enhanced with di-2-ethylhexyl phthalate exposure, and this relationship depended on both the expression levels of the pregnane X receptor as well as concentrations of glucocorticoids^34^. Given that cytochrome p450 enzymes are involved in metabolism of xenobiotics, they may be sensitive to phthalate exposures. Furthermore, since cytochrome p450 enzymes also metabolize endogenous signaling molecules, an increased burden in phthalate exposure may impose adverse perturbations in the normal functions of these enzymes.

Studies that have assessed mediation pathways for phthalates in the context of pregnancy outcomes focused on limited mediators and did not assess whole biomarker groups^35–37^. In the larger LIFECODES cohort, Ferguson and colleagues conducted mediation analysis using the oxidative stress biomarker 8-IP as a potential mediator in the relationship between phthalate metabolites and preterm birth (n_cases_ = 130, n_controls_ = 352)^37^. This study demonstrated that 8-IP mediated the relationship between preterm birth and MEHP, MECPP, and mono-n-butyl phthalate (MBP)^37^. A pilot study based in China (n = 115) investigated DNA methylation signatures as potential mediators but did not find evidence of mediation in the relationship between prenatal phthalates and gestational age^35^. Extending to outcomes beyond pregnancy, a study based in Norway investigated maternal thyroid hormones as potential mediators in the relationship between prenatal phthalate exposure and attention-deficit hyperactivity disorder (n_cases_ = 297, n_controls_ = 553), however there was no evidence of mediation^38^. The lack of studies focused on high-dimensional mediation pathways in the relationships between toxicant mixtures and pregnancy outcomes signifies a major knowledge gap in environmental epidemiology research. Identification of high-dimensional mediation pathways helps prioritize biomonitoring efforts for early interventions. By further validating the mediation effect of cytochrome p450 pathway in independent samples, we may potentially be able to utilize eicosanoid biomarkers of this pathway as early signals of disease states in pregnancy attribute to phthalate exposure.

There are features of our study that can be improved upon in order to advance knowledge of biological pathways that may mediate phthalate induced pregnancy outcomes. Our study was not only limited in sample size, but also by a single cross-sectional assessment of exogenous and endogenous biomarkers. One previous study demonstrated significant changes in the expression of cytochrome p450 enzymes across pregnancy in placental tissue^39^. Another study has also shown that fetal expression levels of cytochrome p450 enzymes vary significantly from gestation to neonatal years^27^. Additionally, the cross-sectional design of our study also underlines potential for reverse causation. For example, elevated inflammatory disease states from other causative agents may be linked to increased cytochrome p450 products, in turn impacting renal clearance of phthalate metabolites measured in urine samples. Future studies should build upon these findings to investigate cytochrome p450 products as mediators in a longitudinal design. Additionally, dimension reduction and shrinkage methodology was limited to a continuous outcome variable, and future innovations in statistical modeling will need to be developed in order to adapt our high-dimensional mediation pipeline for case-control settings with preterm birth. Another limitation in our study is the lack of potential confounders that may contribute to residual confounding. Specifically, dietary information may be an important confounder, given that some toxicants are associated with the type of food products that are consumed.

Our study contains multiple strengths that underline its utility in epidemiologic research. We applied an extensive ensemble of statistical methods in our analytical pipeline. By doing so, we build upon the methodological advancements in high-dimensional mediation by incorporating mixtures analysis – a critical and necessary pre-requisite for understanding the health consequences of the human exposome. We created a guided template for mediation analysis that is widely applicable in various settings that is flexible to different structures of exposure data, mediator biomarkers, and health outcomes. Another strength of our study was the robust exposure assessment that quantified over four different classes of environmental agents that are known or suspected reproductive toxicants. Similarly, we measured a comprehensive panel of eicosanoids, which informs potential biomarkers for health studies.

In conclusion, this study integrates environmental mixtures analysis with cutting edge high-dimensional mediation approaches. Toxicants with sparse effects exhibited subtle mediation effects. However, cumulative exposures, assessed through construction of the phthalate risk score, exhibited evidence for mediation through the cytochrome p450 pathway in the context of decreased gestational age at delivery. In order to accurately estimate the impact of the human exposome, future studies should consider implementation of high-dimensional mediation analysis combined with exposure mixtures methods.

## 4. Methods

### 4.1 Study population

The LIFECODES pregnancy cohort was developed at the Brigham and Women’s Hospital in Boston, MA. The overall cohort of 1,600 pregnant women was recruited between 2006 and 2008. The Brigham and Women’s Hospital administered institutional review board approval for this study. This study sample includes 161 women with complete exposure and endogenous biomarker measurements. Participants were over the age of 18 and recruited early in pregnancy (< 15 weeks gestation). In total, this sample includes 52 cases of preterm birth (defined as delivery < 37 weeks gestation), and 109 controls. Spontaneous preterm birth (n_cases_ = 30, n_controls_ = 109) was defined as either having a preterm premature rupture of the membranes, or spontaneous preterm labor^40^. Questionnaires and physical exams were administered at study visits to collect data on key covariates (e.g. maternal age, race, education, body mass index). Participants provided biological specimens (urine and blood) at a clinic visit occurring between 23.1 and 28.9 weeks gestation. Maternal urine samples were collected at median 26 weeks gestation and subsequently frozen at −80° C. Plasma was collected from 10mL of blood draw using ethylenediaminetetraacetic acid plasma tubes. After collection, blood was stored at +4° C for less than four hours and subsequently centrifuged for 20 minutes before being stored at −80°C. Additional details regarding recruitment and study design for the LIFECODES cohort have been published previously^41,42^.

### 4.2 Exposure analytes

All exposure analytes were measured in urine samples. Quantification of individual toxicants and analytes followed protocols developed by the Centers for Disease Control and Prevention. We used high performance liquid chromatography-electrospray ionization-tandem mass spectrometry (HPLC-ESI-MS/MS) to quantify a total of nine phthalate metabolites. We used isotope dilution liquid chromatography with tandem mass spectrometry (ID-LC-MS/MS) to quantify a total of eight polycyclic aromatic hydrocarbon metabolites and ten phenolic analytes. Finally, a total of 17 trace metals were measured using a Thermo Fisher (Waltham, MA, USA) iCAP RQ inductively coupled plasma mass spectrometer (ICPMS) with Teledyne CETAC Technologies (Omaha, NE, USA) ASX-520 autosampler. Additional details on protocols for exposure assessment have been previously described^43–46^.

### 4.3 Endogenous biomarkers

A large panel of 53 eicosanoids and lipid metabolites were measured in plasma samples using a 6490 Triple Quadrupole mass spectrometer (Agilent, New Castle, DE, USA). The individual eicosanoids were identified using metabolite-specific fragmentation and retention times, and instrumentation parameters have been previously documented^12^. We used the Milliplex MAP High Sensitivity Human Cytokine Magnetic Bead Panel (EMD Millipore Corp., St. Charles, MO) to measure four cytokines: IL-1β, IL-6, TNF-α, and IL-10. An additional inflammation marker, CRP, was also measured using a DuoSet enzyme-linked immunosorbent assay (ELISA) (R&D Systems, Minneapolis, MN). Additional analytical details for measurement of inflammatory markers have been previously described^10^.

We measured three protein oxidation markers in plasma samples: 3-nitrotyrosine [NY], 3-chlorotyrosine [CY], and *o,o’*-dityrosine [DY]. To quantify these biomarkers, total plasma protein was first precipitated and diluted with a phosphate buffer. The samples were then delipidated, injected with isotopically labeled standards, and hydrolyzed for 24 hours. Subsequently, the processed plasma samples underwent liquid chromatography electrospray ionization tandem mass spectrometry. Additional oxidative stress markers 8-IP and 8-OHdG were measured in urine samples at Cayman Chemical (Ann Arbor, MI). 8-IP was quantified using affinity column chromatography followed by an enzyme immunoassay, and 8-OHdG was measured directly through an enzyme immunoassay. Further analytical details for oxidative stress markers have been previously described^47^.

### 4.4 Statistical analyses

Descriptive statistics were weighted based on calculated inverse probability weights that corrected for over-representation of preterm birth cases in the sample compared to the larger LIFECODES cohort.

In **Figure 1**, we illustrate a step-wise guide to the analytical pipeline that we developed for this high-dimensional mediation analysis.

#### Step 1: Pairwise mediation of exposures and endogenous biomarkers

Our analytical pipeline starts with one-at-a-time pairwise mediation analysis with outcome *Y* and every possible unique combination of analytes from the exposure matrix, ***A*** (*p* = 38, where *p* is the number of analytes), and endogenous biomarkers from the mediator matrix, ***M*** (*q* = 63, where *q* is the number of mediators). This results in a total of 2,394 unique mediation models. The goal of this initial step is to conduct an exhaustive screen of all possible combinations of mediation pathways between each toxicant and endogenous biomarker measured in our study. The general framework for the mediation models is represented by:

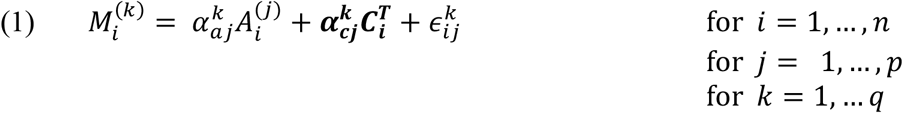

Here *(i, j, k)* stands for the *i*-th subject, *j*-th exposure and *k*-th mediator. In these models, 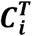, is a covariate vector of *r*-by-*1* (*r* = 5, where the first element is scalar 1 for the intercept and the remainder elements are covariates) for the i-th subject. The individual mediators are log-transformed and denoted by 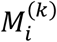 for the *i*-th subject and *k*-th mediator, and the exposure variables are log-transformed and denoted by 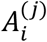 for the *j*-th exposure. In models where gestational age at delivery was the outcome variable, the conditional model for ***Y*** |***A, M, C*** (is modeled as:

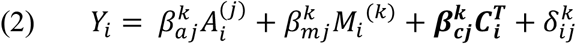

To account for the nested case-control sampling approach, we constructed inverse probability weights so that the subset sample was more representative of the proportion of cases and controls in the larger LIFECDOES cohort. Models in equations 1 and 2 were weighted by these inverse probability weights. The natural indirect effect for each exposure-mediator pair *(j, k)* is estimated by taking the product, 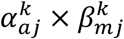. When the conditional model for ***Y*** |***A, M, C***, modeled the binary outcome variables preterm birth and spontaneous preterm birth, the model specification was:

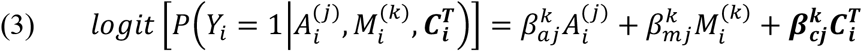

#### Step 2: Multiple mediators: shrinkage and dimension reduction

The second step of our analytical pipeline provides options for evaluating the *q* × *q* mediator matrix in a high-dimensional setting by either conducting shrinkage estimation or reducing the dimensionality of the mediator matrix. For shrinkage estimation, we applied Bayesian shrinkage estimation. In this setting, we deployed a high-dimensional shrinkage mediation analysis, one-at-a-time, for each unique exposure analyte (*p* = 38) in the exposure matrix, ***A*** (**Figure 1**). The major goal of Bayesian shrinkage estimation is to evaluate global mediation effects of multiple mediators simultaneously and extract posterior inclusion probability from a mixture prior as measures of importance among mediation pathways. For each analyte in the exposure matrix, ***A***, we implemented shrinkage estimation by applying Bayesian mediation analysis with the *bama* package (version 0.9.1)^20^. In this application, *bama* modifies equation 1 into a multivariate regression model that jointly evaluates all mediators simultaneously in association with individual toxicants:

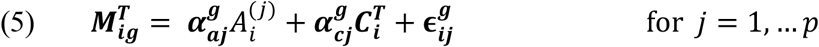

The 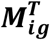 denotes the mediator vector with a length equivalent to the number of mediators which are partitioned into groups *g* (*g* = 1,2,3,4,5,6,7) corresponding to literature derived mediator groups (cyclooxygenase pathway [*q*_1_ = 14], cytochrome p450 pathway [*1*_2_ = 18], lipoxygenase pathway [*q*_5_ = 14], parent lipid compounds [*q*_4_ = 5], inflammation biomarkers [*q*_5_ = 5], oxidative stress biomarkers [*q*_6_ = 2], and protein damage biomarkers [*q*_7_ = 3]). In this setting, 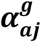 are the *q*_g_ × 1 vectors of coefficients, and 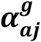 are matrices of dimension *q*_g_ × *r*. Implementation of *bama* is restricted to continuous outcome variables; therefore we focus on gestational age at delivery and modify the model in equation 2 to contain the entire mediator matrix:

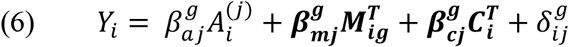

Here 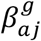 is scalar, representing the estimated coefficient for each exposure given a mediator group, and 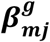 is a 1-by-*q*_g_ vector of coefficients, where *q*_g_ is equal to the number of mediators that belong in each group listed previously. In alignment with the continuous shrinkage approach used in Bayesian sparse linear mixed models^48^, *bama* assumes that the mediation effects are predominantly sparse, with selected active mediators having larger mediation effects. *Bama* utilizes the *L2* norm^49^ to perform shrinkage and selection on the effects of a 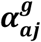 and 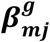 (separately, and for exposure *j* estimates the natural indirect effect as the sum of the products of the individual effects 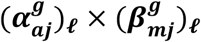 and defines global pathway specific mediation effects as:

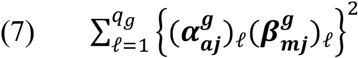

where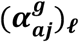 and 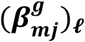 are the *ℓ*-th entries of the vector 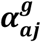 and 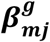, respectively. Prior specification for 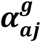 and 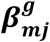 is set to default settings in *bama*, which has been explained previously^20^.

The secondary approach in *step 2* of the analytical pipeline is mediator dimension reduction. The main goal of reducing the dimensionality of the mediator matrix was to determine the extent to which individual toxicants are mediated by whole groups of endogenous biomarkers (cyclooxygenase pathway, cytochrome p450 pathway, lipoxygenase pathway, parent lipid compounds, inflammation biomarkers, oxidative stress biomarkers, and protein damage biomarkers). Specifically, we constructed directions of mediation, ***w***, using population value decomposition^19^. In this application, mediators are again organized into seven groups, *g*. Each mediator within group *g* contributes to the group-specific 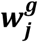 under the consideration of exposure *j*, thus when using the first direction of mediation for each group, we reduce the total dimension of the mediator matrix (from *q* = 63 individual mediators to *s* = 7 groups). The direction of mediation, 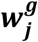, is the coefficient vector of the linear combination of mediators in each group. Each 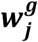 is estimated by maximizing the likelihood of structural equation modeling consisting of the models specified in equations 5 and 6. The coefficients estimated in the first direction of mediation, 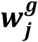, are then used as weights to estimate a mediator group effect, 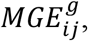 by taking the linear combination of the coefficient weights across each measured mediator for each individual:

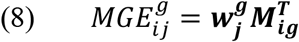

For each exposure analyte in the exposure matrix, ***A***, we create an *MGE* for each mediator group (cyclooxygenase pathway, cytochrome p450 pathway, lipoxygenase pathway, parent lipid compounds, inflammation biomarkers, oxidative stress biomarkers, and protein damage biomarkers), and conduct pair-wise single mediation, 3*-by-*Z, totaling to 266 unique mediation models. The models are specified as:

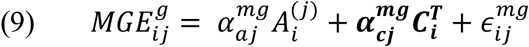

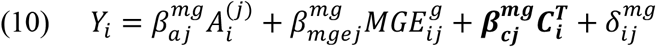

This mediation approach continues to focus strictly on gestational age at delivery, and we estimate the natural indirect effect by taking the product 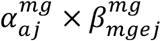.

#### Step 3: Both exposure and mediator dimension reduction

In the third step of our analytical pipeline we introduce a method to construct environmental risk scores^23^, thus reducing the dimensionality of the exposure matrix of dimension 38. The goals in this final step are to estimate the cumulative effect of toxicants based on their respective class, *m* (phthalates [*p*_1_ = 9], phenols [*p*_2_ = 9], polycyclic aromatic hydrocarbons [*p*_3_ = 8], and metals [*p*_4_ = 12]), and then estimate class-level mediation pathways with whole mediator groups (cyclooxygenase pathway, cytochrome p450 pathway, lipoxygenase pathway, parent lipid compounds, inflammation biomarkers, oxidative stress biomarkers, and protein damage biomarkers). In this setting, we reduce the total number of pairwise mediation models to 28. We first estimate weights to construct environmental risk scores by applying adaptive elastic net regularization on toxicants based on their classes:

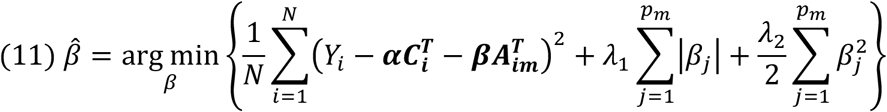

Adaptive elastic net combines the least absolute shrinkage and selection operator penalty with ridge regression^50^. In this setting, *Y_i_* is the continuous outcome variable gestational age at delivery, *p_m_* is the number of exposure analytes in each toxicant class, and 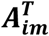 is the vector of, *p_m_* log-transformed and standardized exposures for individual *i*(*i* = 1,…, *n*). In this application, λ_1_ is a tuning parameter that performs shrinkage of coefficients for each exposure variable and λ_2_ operates as a tuning parameter used to stabilize solutions paths that navigate multi-collinearity in the exposure matrix. We utilized 5-fold cross-validations and optimization of prediction errors in order to estimate λ_1_ and λ_2_.

To construct environmental risk scores for each toxicant class, we first extract coefficients estimated by adaptive elastic net and designate them as a vector of weights. We then estimate each individual’s scores, *ERS_im_*, by calculating the linear combination using weights produced from adaptive elastic net, applied across each individual’s observed measurements of each toxicant:

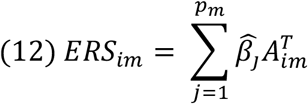

Next, we combine the population value decomposition approach for mediator dimension reduction^19^ and reconstruct an 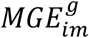 for each *ERS_im_*. Since the *ERS_im_* are standardized, mediators are also log-transformed and standardized in this setting. Thus, we conduct pair-wise mediation analysis for each *ERS_im_* (*v* = 4 risk scores) with each 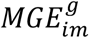 (*s* = 7 groups), resulting in a final reduced combination of 28 mediation models:

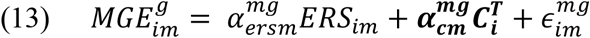

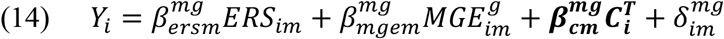

Finally, using each *ERS*, we also applied two additional multivariate mediation methods: (1) pathway lasso^22^ and, (2) high-dimensional mediation analysis using a joint significance test for mediation effect^21^. These methods evaluate specific pairs of \mn and mediators to compare and differentiate group-level findings from sparse mediation effects of individual mediators. We adapt equations 13 and 14 and substitute 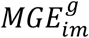with 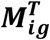 specified in equations 5 and 6:

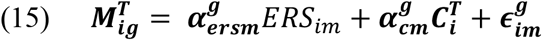

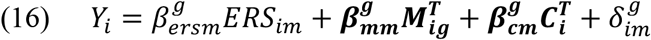

Pathway lasso introduces a novel penalty on the product of indirect effects and performs shrinkage and selection to identify pairs of \mn and single mediators that have the most robust mediation pathway.

We applied the joint significance test for high-dimensional mediation analysis using the *hima* package (version 1.0.7). *Hima* applies the minimax concave penalty (MCP) to regularize the coefficients estimated in equations 15 and 16 and identify important mediating pathways^21^. Subsequently, *hima* conducts a joint significance test based on the MCP-penalized coefficients, resulting in adjusted p-values accounting for multiple comparisons using either the Benjamini Hochberg or Bonferroni method.

## Data Availability

Due to the sensitive nature of biological samples and demographic variables of the human subjects dataset, the data used for this manuscript may not be publicly shared.

## Software

Analytical pipeline available in the form of R scripts available at https://github.com/umich-cphds/environmental_mediation_pipeline.

## Author Contributions

M.A contributed to data pre-processing, data analysis, writing, and interpretation. Y.S. contributed to data analysis, writing, and interpretation. K.F. and D.C. contributed to data pre-processing and interpretation. L.Z. and S.P. contributed to eicosanoid measurement and quantification. J.M., T.M., and B.M. contributed to study design and interpretation.

## Acknowledgements

Subject recruitment and sample collection was originally funded by Abbott Diagnostics. This work was also supported by National Institute of Environmental Health Sciences, National Institutes of Health (grants R01ES018872, P42ES017198, P30ES017885, P50ES026049, and U2CES026553). Support for Kelly Ferguson was provided in part by the Intramural Research Program of the NIH, National Institute of Environmental Health Sciences (ZIAES103321).

## References

1. Schettler, T. Human exposure to phthalates via consumer products. Int J Androl 29, 134– 9– discussion 181–5 (2006).

2. Andra, S. S., Charisiadis, P., Arora, M., van Vliet-Ostaptchouk, J. V. & Makris, K. C. Biomonitoring of human exposures to chlorinated derivatives and structural analogs of bisphenol A. Environment International 85, 352–379 (2015).

3. Błędzka, D., Gromadzińska, J. & Wąsowicz, W. Parabens. From environmental studies to human health. Environment International 67, 27–42 (2014).

4. Alegbeleye, O. O., Opeolu, B. O. & Jackson, V. A. Polycyclic Aromatic Hydrocarbons: A Critical Review of Environmental Occurrence and Bioremediation. Environmental Management 1–26 (2017). doi:10.1007/s00267-017-0896-2

5. Singh, A. & Prasad, S. M. Remediation of heavy metal contaminated ecosystem: an overview on technology advancement. Int. J. Environ. Sci. Technol. 12, 353–366 (2014).

6. Vrijheid, M., Casas, M., Gascon, M., Valvi, D. & Nieuwenhuijsen, M. Environmental pollutants and child health—A review of recent concerns. International Journal of Hygiene and Environmental Health 219, 331–342 (2016).

7. Kiyama, R. & Wada-Kiyama, Y. Estrogenic endocrine disruptors: Molecular mechanisms of action. Environment International 83, 11–40 (2015).

8. Milnerowicz, H., Ściskalska, M. & Dul, M. Pro-inflammatory effects of metals in persons and animals exposed to tobacco smoke. Journal of Trace Elements in Medicine and Biology 29, 1–10 (2015).

9. Ferguson, K. K. et al. Repeated measures of urinary oxidative stress biomarkers during pregnancy and preterm birth. The American Journal of Obstetrics & Gynecology 212,208.e1–208.e8 (2015).

10. Ferguson, K. K., McElrath, T. F., Chen, Y.-H., Mukherjee, B. & Meeker, J. D. Longitudinal Profiling of Inflammatory Cytokines and C-reactive Protein during Uncomplicated and Preterm Pregnancy. Am J Reprod Immunol 72, 326–336 (2014).

11. Aung, M. T. et al. Prediction and associations of preterm birth and its subtypes with eicosanoid enzymatic pathways and inflammatory markers. Scientific Reports 9, 17049– 17 (2019).

12. Afshinnia, F. et al. Elevated lipoxygenase and cytochrome P450 products predict progression of chronic kidney disease. Nephrology Dialysis Transplantation 1359, 60–10 (2018).

13. Spector, A. A., Fang, X., Snyder, G. D. & Weintraub, N. L. Epoxyeicosatrienoic acids (EETs): metabolism and biochemical function. Progress in Lipid Research 43, 55–90 (2004).

14. Dalle Vedove, F. et al. Increased epoxyeicosatrienoic acids and reduced soluble epoxide hydrolase expression in the preeclamptic placenta. Journal of Hypertension 34, 1364–1370 (2016).

15. Ferguson, K. K., McElrath, T. F. & Meeker, J. D. Environmental phthalate exposure and preterm birth. JAMA Pediatr 168, 61–67 (2014).

16. Boss, J. et al. Associations between mixtures of urinary phthalate metabolites with gestational age at delivery: a time to event analysis using summative phthalate risk scores. Environmental Health 17, 56–13 (2018).

17. Baron, R. M. & Kenny, D. A. The moderator-mediator variable distinction in social psychological research: conceptual, strategic, and statistical considerations. J Pers Soc Psychol 51, 1173–1182 (1986).

18. Rubin, D. B. Estimating causal effects of treatments in randomized and nonrandomized studies. Journal of educational Psychology 66, 688 (1974).

19. Chén, O. Y. et al. High-dimensional multivariate mediation with application to neuroimaging data. Biostatistics 19, 121–136 (2017).

20. Song, Y. et al. Bayesian shrinkage estimation of high dimensional causal mediation effects in omics studies. Biom biom.13189–11 (2019). doi:10.1111/biom.13189

21. Zhang, H. et al. Estimating and testing high-dimensional mediation effects in epigenetic studies. Bioinformatics 32, 3150–3154 (2016).

22. Zhao, Y. & Luo, X. Pathway Lasso: Estimate and Select Sparse Mediation Pathways with High Dimensional Mediators. (2016).

23. Park, S. K., Zhao, Z. & Mukherjee, B. Construction of environmental risk score beyond standard linear models using machine learning methods: application to metal mixtures, oxidative stress and cardiovascular disease in NHANES. 1–17 (2017). doi:10.1186/s12940-017-0310-9

24. Aung, M. T. et al. Manganese is associated with increased plasma interleukin-1β during pregnancy, within a mixtures analysis framework of urinary trace metals. Reprod. Toxicol. 93, 43–53 (2019).

25. Aung, M. T., Ferguson, K. K., Cantonwine, D. E., McElrath, T. F. & Meeker, J. D. Preterm birth in relation to the bisphenol A replacement, bisphenol S, and other phenols and parabens. Environmental Research 169, 131–138 (2018).

26. Kim, S. S. et al. Urinary trace metals individually and in mixtures in association with preterm birth. Environment International 121, 582–590 (2018).

27. Sadler, N. C. et al. Hepatic Cytochrome P450 Activity, Abundance, and Expression Throughout Human Development. Drug Metabolism and Disposition 44, 984–991 (2016).

28. Deng, Y., Theken, K. N. & Lee, C. R. Cytochrome P450 epoxygenases, soluble epoxide hydrolase, and the regulation of cardiovascular inflammation. Journal of Molecular and Cellular Cardiology 48, 331–341 (2010).

29. Carroll, M. A. et al. Cytochrome P-450-dependent HETEs: profile of biological activity and stimulation by vasoactive peptides. Am. J. Physiol. 271, R863–9 (1996).

30. Powell, W. S. & Rokach, J. Biosynthesis, biological effects, and receptors of hydroxyeicosatetraenoic acids (HETEs) and oxoeicosatetraenoic acids (oxo-ETEs) derived from arachidonic acid. BBA – Molecular and Cell Biology of Lipids 1851, 340– 355 (2015).

31. Ferguson, K. K. & Chin, H. B. Environmental chemicals and preterm birth: Biological mechanisms and the state of the science. Curr Epidemiol Rep 4, 56–71 (2017).

32. Romero, R. et al. A Role for the Inflammasome in Spontaneous Labor at Term. Am J Reprod Immunol 11, n/a–n/a (2016).

33. Ozaki, H., Sugihara, K., Watanabe, Y., Ohta, S. & Kitamura, S. Cytochrome P450-inhibitory activity of parabens and phthalates used in consumer products. J Toxicol Sci 41, 551–560 (2016).

34. Cooper, B. W., Cho, T. M., Thompson, P. M. & Wallace, A. D. Phthalate induction of CYP3A4 is dependent on glucocorticoid regulation of PXR expression. Toxicological Sciences 103, 268–277 (2008).

35. Huang, L.-L. et al. Prenatal phthalate exposure, birth outcomes and DNA methylation of Alu and LINE-1 repetitive elements: A pilot study in China. Chemosphere 206, 759–765 (2018).

36. Engel, S. M. et al. Prenatal Phthalates, Maternal Thyroid Function, and Risk of Attention-Deficit Hyperactivity Disorder in the Norwegian Mother and Child Cohort. Environ Health Perspect 126, 057004–11 (2018).

37. Ferguson, K. K. et al. Mediation of the Relationship between Maternal Phthalate Exposure and Preterm Birth by Oxidative Stress with Repeated Measurements across Pregnancy. Environ Health Perspect 125, 488–494 (2017).

38. Engel, S. M. et al. Prenatal Phthalate Exposure Is Associated with Childhood Behavior and Executive Functioning. Environ Health Perspect 118, 565–571 (2010).

39. Cizkova, K. & Tauber, Z. Time-dependent expression pattern of cytochrome P450 epoxygenases and soluble epoxide hydrolase in normal human placenta. Acta Histochem. 120, 513–519 (2018).

40. McElrath, T. F. et al. Pregnancy Disorders That Lead to Delivery Before the 28th Week of Gestation: An Epidemiologic Approach to Classification. American Journal of Epidemiology 168, 980–989 (2008).

41. McElrath, T. F. et al. Longitudinal evaluation of predictive value for preeclampsia of circulating angiogenic factors through pregnancy. The American Journal of Obstetrics & Gynecology 207, 407.e1–407.e7 (2012).

42. Ferguson, K. K., McElrath, T. F., Ko, Y.-A., Mukherjee, B. & Meeker, J. D. Variability in urinary phthalate metabolite levels across pregnancy and sensitive windows of exposure for the risk of preterm birth. Environment International 70, 118–124 (2014).

43. Ferguson, K. K., McElrath, T. F., Mukherjee, B., Loch-Caruso, R. & Meeker, J. D. Associations between Maternal Biomarkers of Phthalate Exposure and Inflammation Using Repeated Measurements across Pregnancy. PLoS ONE 10, e0135601 (2015).

44. Ferguson, K. K. et al. Urinary Polycyclic Aromatic Hydrocarbon Metabolite Associations with Biomarkers of Inflammation, Angiogenesis, and Oxidative Stress in Pregnant Women. Environ. Sci. Technol. 51, 4652–4660 (2017).

45. Aung, M. T. et al. Associations between maternal plasma measurements of inflammatory markers and urinary levels of phenols and parabens during pregnancy: A repeated measures study. Science of the Total Environment, The 650, 1–10 (2019).

46. Kim, S. S. et al. Urinary trace metals individually and in mixtures in association with preterm birth. Environment International 121, 582–590 (2018).

47. Ferguson, K. K. et al. Urinary Polycyclic Aromatic Hydrocarbon Metabolite Associations with Biomarkers of Inflammation, Angiogenesis, and Oxidative Stress in Pregnant Women. Environ. Sci. Technol. acs.est.7b01252–9 (2017). doi:10.1021/acs.est.7b01252

48. Zhou, X., Carbonetto, P. & Stephens, M. Polygenic modeling with bayesian sparse linear mixed models. PLOS Genetics 9, e1003264 (2013).

49. Huang, Y.-T. & Pan, W.-C. Hypothesis test of mediation effect in causal mediation model with high-dimensional continuous mediators. Biom 72, 402–413 (2015).

50. Zou, H. & Zhang, H. H. On the adaptive elastic-net with a diverging number of parameters. Ann. Statist. 37, 1733–1751 (2009).

